# Parental feeding and childhood genetic risk for obesity: Exploring hypothetical interventions with causal inference methods

**DOI:** 10.1101/2021.01.07.21249377

**Authors:** Moritz Herle, Andrew Pickles, Nadia Micali, Mohamed Abdulkadir, Bianca De Stavola

**Affiliations:** Department of Biostatistics & Health Informatics, Institute of Psychology, Psychiatry and Neuroscience, Kings College London, UK; Department of Pediatrics Gynaecology and Obstetrics, Faculty of Medicine, University of Geneva, Geneva, Switzerland; Department of Psychiatry, Faculty of Medicine, University of Geneva, Geneva, Switzerland; Population, Policy & Practice, UCL Great Ormond Street Institute of Child Health, University College London, London, UK

**Keywords:** ALSPAC, causal inference, childhood obesity, parental feeding, polygenic risk score

## Abstract

Parental feeding behaviors are common intervention targets for childhood obesity, but often only deliver small changes. Childhood BMI is partly driven by genetic effects, and the extent to which parental feeding interventions can mediate child genetic liability is not known. Here we aim to examine how potential interventions on parental feeding behaviors can mitigate some of the association between child genetic liability and BMI in early adolescence, using causal inference based methods. Data were from the Avon Longitudinal Study of Parents and Children and we quantified the interventional disparity measure of child genetic risk for BMI (PRS-BMI) on objectively BMI at 12 years, if we were to intervene on parental feeding styles measured when children were 10-11 years (n=4,248). Results are presented as Adjusted Total Association (Adj-Ta) between genetic liability and BMI at 12 years, versus the Interventional Disparity Measure Direct Effect (IDM-DE), which represents the association, that would remain, had we intervened on the parental feeding. For children with the top quintile of genetic liability, an intervention shifting parental feeding to the levels of children with lowest genetic risk, resulted in a difference of 0.81 kg/m^2^ in BMI at 12y (Adj-Ta= 3.27, 95%CI: 3.04, 3.49; versus IDM-DE=2.46, 95%CI: 2.24, 2.67). Findings suggest that parental feeding interventions have the potential to buffer some of the genetic liability for childhood obesity. Further, we highlight a novel way to analyze potential interventions for health conditions only using secondary data analyses, by combining methodology from statistical genetics and social epidemiology.

## Introduction

Childhood obesity remains one of the greatest public health challenges across the globe. The estimated percentage of children and adolescents who meet criteria for obesity (BMI >30 kg/m^2^) has risen from ∼4 to ∼15% between 1975 to 2016 (1). This is especially worrying, as childhood obesity tends to persist into adulthood (2) and child, and adult obesity, have been associated with different negative health outcomes, such as cardiovascular disease (3), depression (4) and asthma (5). Individuals with larger bodies face stigmatization and discrimination which have been found to exacerbate negative health outcomes (6). Specifically, childhood overweight and obesity has consistently found to be associated with greater bullying victimization (7) which can have lifelong health consequences (8). The rate of obesity is higher among children from poorer and marginalized population groups (9). Changes in the food environment, such as increased portion sizes and availability of cheap high energy dense foods, has been highlighted as key drivers for this rapid increase in obesity (10). On a genetic level, recent genome wide association studies have identified >100 genomic markers associated with greater BMI, which when added together have been found to explain ∼6% of variance (11).

Even though biological, and environmental risk factors have been identified, interventions to prevent childhood obesity remain ineffective (12), with randomized control trials (RCT) meta-analyses suggesting that there is some evidence that combined diet and physical activity intervention can result in only small reductions in BMI in younger children (13). Apart from diet and physical activity, some interventions aim to educate and change parents’ (or caregiver’s) behaviors to help them support the children’s growth and nutrition. One specific target of family-based interventions is parental feeding practices, which describe parenting behaviors employed to regulate the child’s food intake and eating behaviors (14). These interventions are based on observational research exploring the association between parental feeding, their children’s eating behaviors and weight (15). RCTs have found some evidence that interventions targeting parental feeding practices resulted in changes in child eating behaviors as well as small decreases in child weight (13, 16, 17).

However, none of these studies have investigated the extent to which genetic liability for obesity might be impacting their effectiveness. This is important, as work by Selzam et al has suggested that parental feeding practices are in part influenced by the child’s genetic liability for obesity. Parents were found to be more likely to restrict their children’s food intake, if their child had greater liability (18). Further, parents’ genetic liability might not only potentially influence their feeding style, but also might be shared with child genetic liability.

Despite the genetic contribution to BMI, and the limited success of parental feeding interventions, there is currently little research investigating the processes linking genetic liability to childhood obesity via parental feeding with the aim of identifying possible interventions along that pathway. However, this would be essential to provide context for the development and evaluation of new potential interventions. Further, mothers’ have reported feeling guilty about passing on genetic propensity for obesity to their children (19), and a greater understanding of this area might help clinicians to alleviate concerns and communicate effectively with parents (20).

In our study, we are combining methods used in health disparity research (21), causal inference mediation analyses (22), and genetic epidemiology (23), to address these research questions using data from a cohort of children born in the southwest of the UK, Avon Longitudinal Study of Parents and Children (ALSPAC). We hypothesize that parental feeding practices during childhood are intermediate factors on the pathway between genetic liability and child BMI. We aim to investigate how potential interventions that change parental feeding practices could mitigate some of the genetic liability for obesity in childhood.

## Results

### Descriptive Statistics

Table 1 compares the distribution of the baseline characteristics of the ALSPAC participants and the subsample analyzed in this study. Regarding the mediators, Supplementary Table 1 lists the responses on the thirteen items probing parental feeding behaviors as well as their subscales (Restriction, pressure to eat, and emotional eating). Supplementary Table 2 shows the mean and standard deviation of the exposure PGS-BMI in the five subgroups defined by the PGS-BMI quintiles. Supplementary Table 3 shows pairwise correlations between exposure, mediators, and outcome. As expected, there is a positive association between PGS-BMI and BMI at 12 years (r=0.36). There is also a positive association between PGS-BMI and restriction (r=0.12), and negative ones between pressure to eat and the other two parental behavior behaviors (−0.22 and −0.23).

**Table 1.** Characteristics of the subsamples of the Avon Longitudinal Study of Parents and Children (ALSPAC) at baseline and analyses sample.

| Characteristics |  | Participants at baseline<br>(alive at one year and<br>part of the original core<br>ALSPAC cohort<br>(n = 13,782) | Participants in analyses sample<br>(complete cases, n= 4,248) |
| --- | --- | --- | --- |
|  |  |  | **Mean (SD) or n (%)** |
| **Sex** | Boys | 7,110 (52%) | 2,096 (49%) |
|  | Girls | 6,672 (48%) | 2,152 (51%) |
| **Maternal education** |  |  |  |
|  | **Less**<br>**than A-**<br>**levels** | 7,916 (65%) | 2,248 (53%) |
|  | **A-levels**<br>**or higher** | 4,330 (35%) | 2,000 (47%) |
| **Maternal BMI**<br>**before pregnancy** |  | 22.9 (3.9), n = 11,391 | 22.8 (3.6) |
| **PGS-BMI** |  | 0.29 (0.28), n=8,654 | 0.27 (0.28) |
| **Latent parental feeding score**<br>**at age 10.7 years, N=7,642** |  |  |  |
| **Emotional feeding** |  | -0.002 (0.48) | -0.02 (0.44) |
| **Restriction** |  | 0.01 (0.51) | -0.01 (0.50) |
| **Pressure to eat** |  | 0.01 (0.63) | 0.00 (0.62) |
| **BMI at 12 years**<br>**(kg/m<sup>2</sup>)** |  | 19.1 (3.4), n=6,651 | 19.0 (3.3) |

The observed distributions of the three latent parental feeding behaviors are illustrated in Figure 2, separated by into genetic liability quintiles. These figures guide the reader through the two hypothetical interventions. In intervention 1 the distributions of parental feeding behaviors are shifted to that of the lowest genetic liability quintile (red lines, Figure 2). In intervention 2, the distributions are shifted “down” one category, for example the distributions of children in liability 5^th^ quintile (pink lines, Figure 2) are shifted to those in the fourth quintile (blue lines, Figure 2).

**Figure 1.**
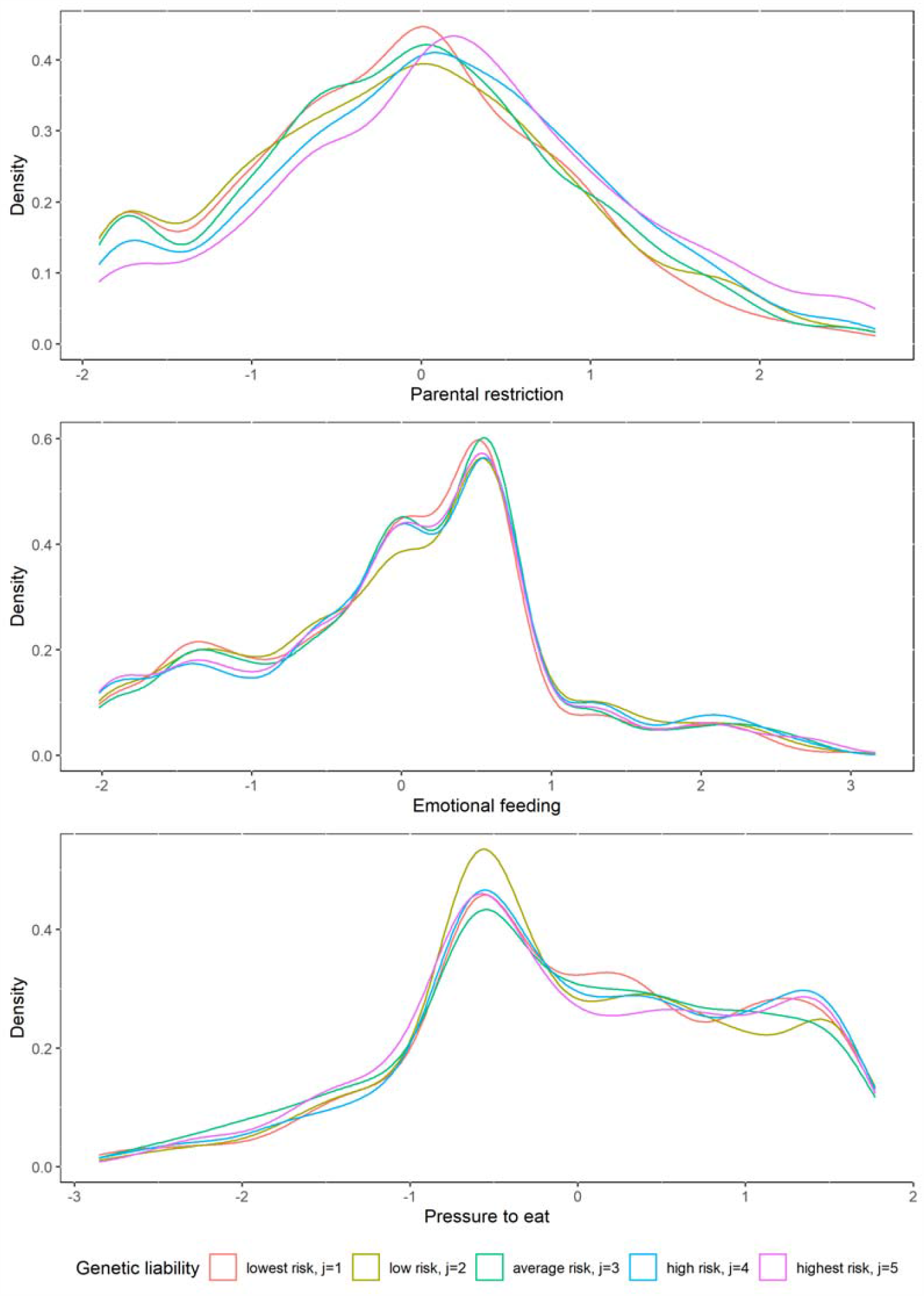
Kernel density plots of parental restriction, emotional feeding and pressure to eat by different quintiles of genetic risk, (j=1,2,3,4,5).

**Figure 2.**
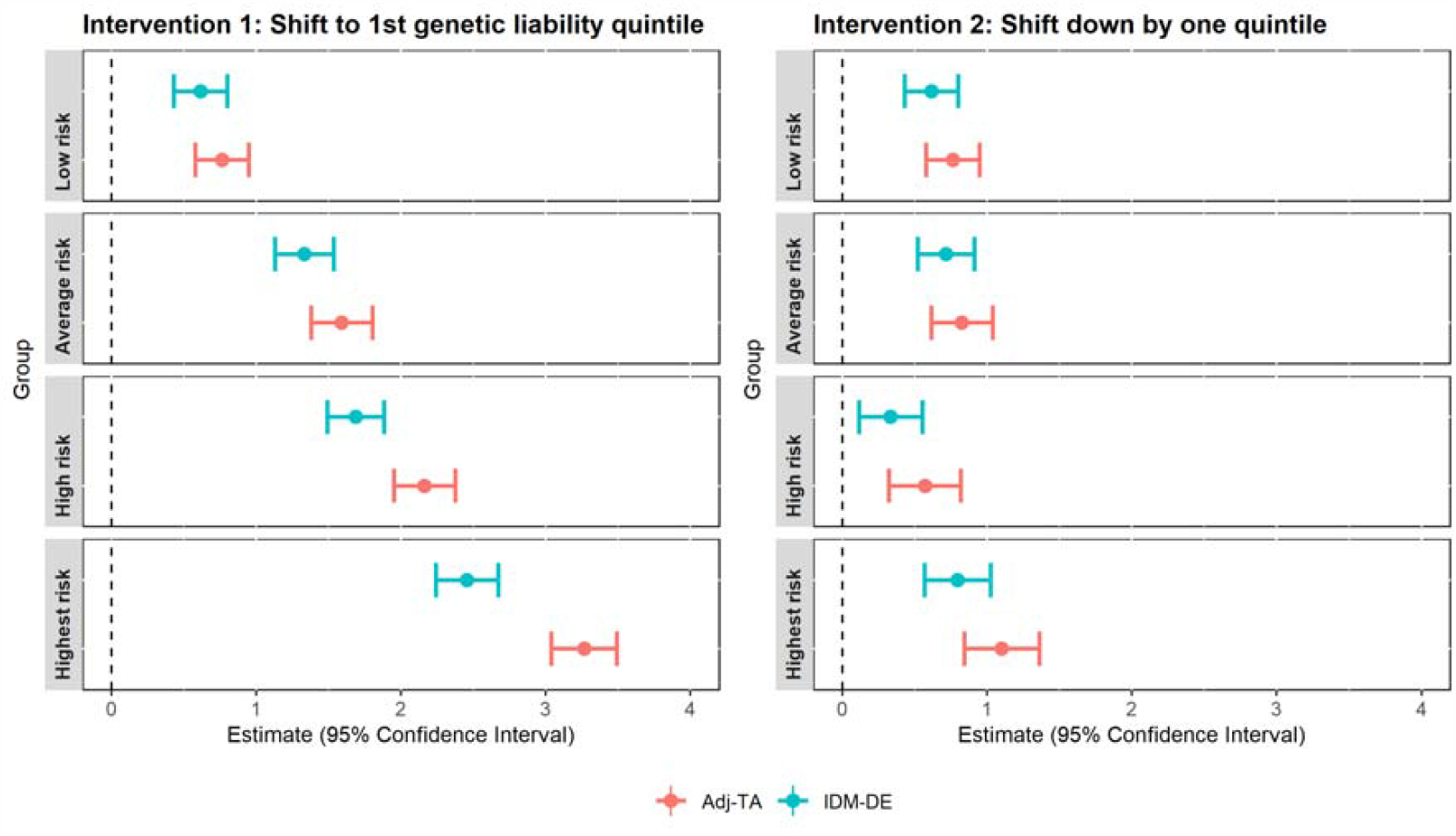
Intervention 1. Adjusted total association (Adj-TA) and Interventional Disparity Measure – Direct Effect (IDM-DE), given a hypothetical intervention shifting the distribution of parental feeding to the distribution under lowest genetic risk (j=1), n= 4,248. Intervention 2. Adjusted total association (Adj-TA) and Interventional Disparity Measure – Direct Effect (IDM-DE), given a hypothetical intervention shifting the distribution of parental feeding to the distribution under average genetic risk (j=3), n= 4,248

### Interventional disparity measures-direct effects

#### Intervention 1: Shifting the distribution of parental feeding to the distribution under lowest genetic liability (j=1)

The first potential intervention shifts the distribution of the three parental feeding behaviors to where they would be in the lowest genetic quintile (j=1). Estimates for IDM-DE and Adj-TA for this setting are presented in Table 2a, and illustrated in Figure 3a. The greatest change in disparity was found for the highest risk quintile (j=5), where the shift in parental feeding resulted in a difference of 0.81 kg/m^2^ (95%CI: 0.67, 0.94) in BMI at 12y (Adj-TA_5_= 3.27, 95%CI: 2.04, 3.49 versus IDM-DE_5_= 2.46, 95%CI: 2.24, 2.67). A smaller difference of 0.47 kg/m^2^ (95% CI: 0.35, 0.59) was found for the high risk 4^th^ quintile (j=4) (Adj-TA_4_ = 2.16, 95%CI: 1.95, 2.38 versus IDM-DE_4_=1.69, 95%CI: 1.49, 1.89). For the 3^rd^ (average liability) and 2^nd^ (low liability) quintiles, IDM-DEs and Adj-TAs had overlapping confidence intervals, indicating little reduction in disparity by the intervention.

**Table 2a.**
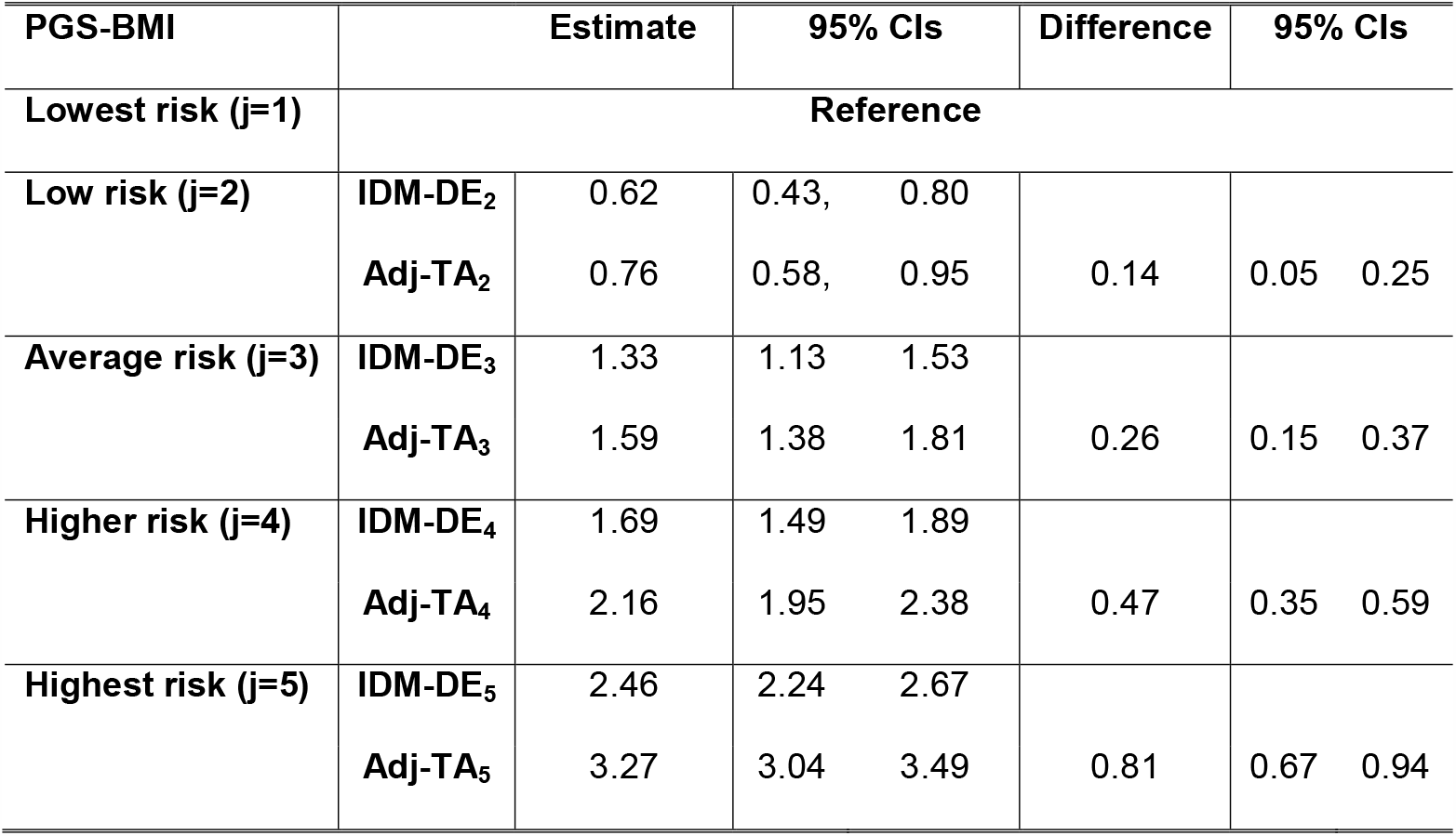
Interventional Disparity Measure – Direct Effect (IDM-DE) and adjusted total association (Adj-TA) of categorical PGS-BMI versus the reference category of average genetic risk (j=1): estimates and 95% Confidence intervals, n= 4,248

**Figure 3.**
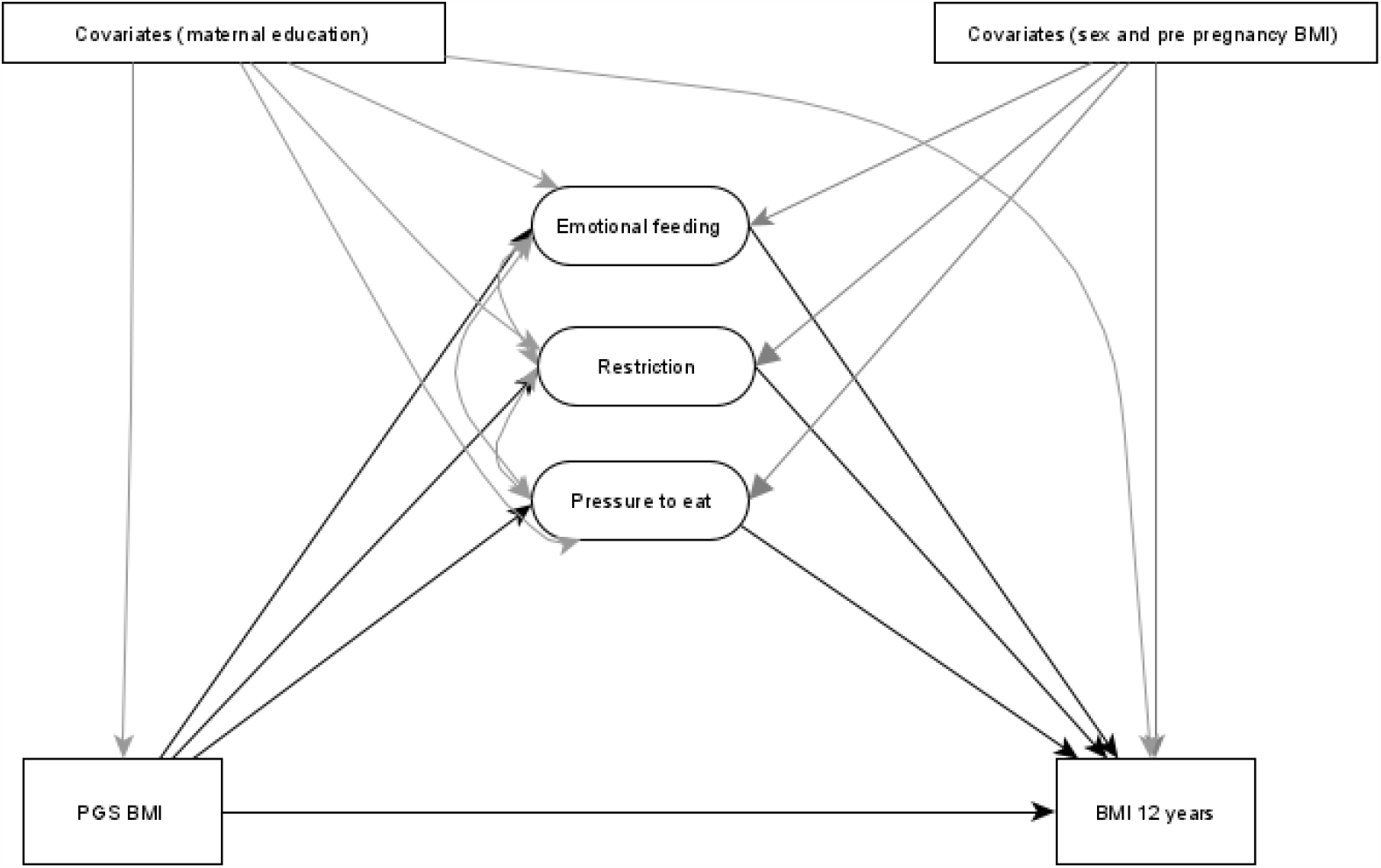
Conceptual model illustrating the associations between exposure (PGS-BMI), mediators (Emotional feeding, restriction, and pressure to eat), outcome (BMI at 12 years) and covariates (sex, pre pregnancy BMI and maternal education).

#### Intervention 2: Shifting down the distribution of parental feeding by one quintile of liability

The second potential intervention shifts the distribution of parental feeding measures to where they would be if the child were one genetic liability quintile lower than they are observed to be. Estimates for Adj-TA, IDM-DE are presented in Table 2b, and illustrated in Figure 3b. For all comparisons, the shift in distributions resulted in moderately smaller IDM-DEs than Adj-TA. The greatest differences were found by shifting from fifth to fourth genetic liability quintiles, from Adj-TA_54_= 1.10 (95% CI: 0.85, 1.36) to IDM-DE_54_ = 0.80 (95%CI: 0.57, 1.02), resulting in a difference of 0.31 (95%CI: 0.17, 0.44). However, confidence intervals were overlapping, as well as for all other comparisons.

**Table 2b.** Interventional Disparity Measure – Direct Effect (IDM-DE) and adjusted total association (Adj-TA) of categorical PGS-BMI if shifted down by one quintile of PGS liability: estimates and 95% Confidence intervals, n= 4,248

| **PGS-BMI** |  | **Estimate** | **95% CIs** |  | **Difference** | **95% CIs** |  |
| --- | --- | --- | --- | --- | --- | --- | --- |
| **Low (j=2)**<br>**to Lowest risk (j=1)** | **IDM-DE<sub>21</sub>** | 0.62 | 0.43 | 0.80 |  |  |  |
|  | **Adj-TA<sub>21</sub>** | 0.76 | 0.58 | 0.95 | 0.14 | 0.05 | 0.25 |
| **Average (j=3)**<br>**to Low risk (j=2)** | **IDM-DE<sub>32</sub>** | 0.72 | 0.52 | 0.91 |  |  |  |
|  | **Adj-TA<sub>32</sub>** | 0.83 | 0.61 | 1.04 | 0.11 | 0.01 | 0.21 |
| **High (j=4)**<br>**to average risk (j=3)** | **IDM-DE<sub>43</sub>** | 0.33 | 0.11 | 0.55 |  |  |  |
|  | **Adj-TA<sub>43</sub>** | 0.57 | 0.32 | 0.82 | 0.24 | 0.12 | 0.36 |
| **Highest (j=5)**<br>**to high risk (j=4))** | **IDM-DE<sub>54</sub>** | 0.80 | 0.57 | 1.02 |  |  |  |
|  | **Adj-TA<sub>54</sub>** | 1.10 | 0.85 | 1.36 | 0.31 | 0.17 | 0.44 |

## Discussion

These results replicate previous work (18), indicating that greater child polygenic liability for obesity is associated with greater parental restriction, marked by the tendency of the parents to control their child’s food intake by, for example, keeping food out of the child’s reach. These findings are in line with previous findings suggesting that to a certain extent parental feeding practices are in response to the child’s weight and hence their genetic liability (24).

Our two potential interventions highlight that changing parental feeding strategies has the capacity to mitigate some of the genetic liability associated with a higher childhood BMI. These findings, if the underlying assumptions are met, support parental feeding strategies as promising intervention targets for child obesity interventions. Previous intervention studies which aimed to change parental feeding strategies have shown some success in changing children’s eating behaviors, associated with later weight, such as decreased responsiveness to external food cues and increased sensitivity to satiety cues (16), as well greater in fruit and vegetable consumption (25) and small reduction of waist circumference (17) and BMI (26).

However, most interventions targeting parental feeding practices have not been able to show significant changes in overall energy intake or measure of body size (16, 27, 28). When comparing our results with previous randomized control trials, it is important to consider that our hypothetical interventions only targeted parental feeding. Most previous interventions have a more holistic approach often including other factors as well, such as sedentary behaviors and emotional regulation. Further, the potential change in parental feeding might not only directly influence child weight but might additionally work through other pathways such as these. For example, Steinsbekk et al have suggested that parental feeding behaviors link with child eating behaviors (29), which in turn influence food intake and child BMI (30, 31). This is of importance, as childhood eating behaviors and childhood body size have been found to share some genetic etiology (32).

Our analyses suggest that potential interventions would be most effective (in absolute terms) for children in the highest quintile of genetic liability. Hence, future interventions might be specifically targeted to families and children at greatest risk of obesity, genetic or otherwise. However, it is important to acknowledge that our study only indexed (common) genetic risk, and families might be vulnerable to obesity due to their socio-economic position and discrimination. Our results only focus on the disparity caused by common genetic differences, included in this polygenic risk score, and more work is necessary to understand these findings in context of wider non-genetic risk factors.

Overall, our findings indicate that changes in parental feedings strategies have the potential to mitigate some of the disparity caused by genetic risk, as they contribute to its association with childhood BMI. This information might be helpful to parents and pediatricians. Previous qualitative research has indicated that parents want conversations with health professionals about risk for obesity and have a great sense of responsibility for protecting their children. Further, parents have acknowledged that conversations around this topic can lead to self-blame and guilt (33). Our new findings might be helpful in this context, as they can highlight that even in the face of increased genetic liability, parental feeding strategies might still make a difference. This would be especially important for vulnerable families who might need support with reflecting on the role of parental feeding and childhood obesity (34).

In addition to implications to parents and pediatricians, we propose a novel direction on how to investigate the potential mitigation of genetic liability by intervening on environmental factors. Drawing form previous work on interventional disparity effects (21, 35), this approach has the ability to employ hypothetical scenarios to tentatively map out what real-life interventions may be able to achieve. Of course, the included models only imperfectly reflect the reality of interventions and etiology of complex health outcomes. Importantly, this approach is pragmatic and cost-effective, as it uses preexisting datasets from longitudinal cohorts, under certain, defensible, assumptions. We believe that this approach could be applied to other health outcomes, which have found to have strong genetic underpinnings such as schizophrenia (36) or coronary heart disease (37).

Over the past years, genetic research has contributed to our understanding of the biology of many health outcomes in children and adults. To move forward, research must go further, and aim to investigate causal questions, drawing data from readily available cohorts. Previous research has implemented instrumental variable approaches, in the form of Mendelian randomization studies, as well as the long history of direction-of-causation twin studies in order to examine the causal direction between variables (38). We believe that the interventional disparity measure approach employed here is a useful addition to the repertoire of analysis tools for researchers studying environmental mediation of genetic risk, linking basic science research with policy.

### Strength & Limitations

The following limitations need to be considered. Parental feeding strategies were measured combining items from the Child Feeding Questionnaire (39) and Parental Feeding Style questionnaire (14) and henceforth cannot be directly compared to other studies, who often only use one or the other psychometric tool. However, results were similar to previous research (18) lending support to these measures. In addition, parents reported their parental feeding behaviors when their children were about 10.7 years old. Many previous interventions targeted parents with younger children, as older children have more autonomy making more of their own choices about what and how much to eat. However, previous research has suggested that parental feeding tracks over time (40), and hence, measures at 10-11 years are likely to reflect earlier parental behaviors. In addition, even though recent genome-wide studies of BMI in multi-ancestry populations have been published (41), our study only included families of European ancestry. Parental feeding practices have been found to differ between ethnic groups in the UK, whereby mothers of South Asian descent reported higher pressure to eat and emotional feeding than mothers from white or black British backgrounds (42). The polygenic risk score included in these analyses is a composite score made from common SNPs and does not include other types of genetic variation such rare variants or coy number variations.

Furthermore, our analyses are based on the assumptions of no interference, consistency, and no unmeasured confounding of mediator-outcome associations. Interference would be present if the parental feeding of one mother would impact the BMI of another child. This seems highly unlikely, as the families in this cohort were recruited from a large region and we included only one child from multiple-sibships. The consistency assumption implies that the value of the intervention target (parental feeding) shifted by hypothetical intervention is the same as the value if it were to be observed. In other words, the observed distribution of emotional feeding for parents with a child of average genetic risk is consistent with the distribution of emotional feeding after it was shifted to the distribution of children with average genetic risk by the hypothetical intervention. This assumption implies that the intervention is “noninvasive” meaning that the outcome for children would not change, if their parents’ feeding behaviors were set to the same value as it was observed. Extensive discussion on this can be found in work by Hernán & VanderWeele (2011) and VanderWeele & Hernán (2013) (43, 44). Regarding unmeasured confounding of the mediator-outcome association, we have included three confounders maternal education, maternal BMI before pregnancy and child sex to capture at least partly some of the confounding.

As mentioned above, our hypothetical interventions only target parental feeding strategies, whereas previous real-life interventions target parental feeding, diet, and physical activity. In our current model, the three included parental feeding practices are considered as joint mediators, and it would be possible to add further potential intervention targets. However, due to the potential complex correlation structure of these additional mediators as well as difficulties around interpretation of findings, we believe that this simpler version is the most appropriate. Further, it can be argued that future interventions by health professionals are easiest to deliver if they target parents and parental behaviors directly, hence these should take priority.

## Conclusions

The results of this study replicate the previously described association between child genetic liability for obesity and parental restrictive feeding practices. Further, our findings suggest that potential interventions targeting parental feeding practices would mitigate some of the disparity caused by genetic liability as measured by a PGS-BMI, especially for children at high risk. These findings emphasize the potential power of interventions aimed at educating and changing in parental feeding practices to give them the tools to support the healthy growth of their children. In addition, by using statistical genetics instruments in the context of causal inference mediation analyses, we propose a novel framework on how to investigate gene-environment interplay when studying complex health outcomes in pediatrics and general health.

## Materials and Methods

### Methods

#### Sample

Participants included in this study are a subsample of adolescents of the population-based ALSPAC cohort that recruited pregnant women in the southwest of England (45, 46). All pregnant women that were expected to have a child in the period of 1 April 1991 until 31 December 1992 were contacted to participate in the original cohort. At the beginning, 14,451 pregnant women took part and 13,988 children were alive at the end of year one. To guarantee independence of individuals, one sibling per set of multiple births (n = 203 sets) is randomly included in our sample. For these analyses, the final subsample included participants who had data on exposure, mediators, and outcome (defined below; n=4,248). Please note that the study website contains details of all the data that are available through a fully searchable data dictionary and variable search tool and reference the following webpage: http://www.bristol.ac.uk/alspac/researchers/our-data/.

Ethical approval for the ALSPAC participants was obtained from the ALSPAC Ethics and Law Committee and the Local Research Ethics Committees: www.bristol.ac.uk/alspac/researchers/research-ethics/. Consent for biological samples was collected in accordance with the Human Tissue Act (2004).

#### Measures

##### Exposure

Genotype data were available for 9915 children out of the total of 15,247 ALSPAC participants. Participants were genotyped on the genome-wide Illumina HumanHap550 quad chip. Individuals with disproportionate levels of individual missingness (i.e., >3%), insufficient sample replication (identity by descent < 0.8), biological sex mismatch, and non-European ancestry (as defined by multi-dimensional scaling using the HapMap Phase II, release 22, reference populations) were excluded. SNPs with a minor allele frequency (MAF) of < 1%, excessive missingness (i.e., call rate < 95%), or a departure from the Hardy–Weinberg equilibrium (P value < 5 × 10^−7^) were removed. Imputation was conducted with Impute3 using the HRC 1.0 as the reference panel (47) and phasing was carried out using ShapeIT (v2.r644). Finally, post-imputation quality control checks were performed; any SNPs with MAF less than 1%, Impute3 information quality metric of < 0.8, and not confirming to Hardy-Weinberg equilibrium (P < 5 × 10-7) were removed. After data cleaning, a total of 8,654 individuals and 4,054,653 SNPs remained eligible for analyses.

Polygenic scores (PGS) were derived from summary statistics of the Genetic Investigation of Anthropometric Traits (GIANT) consortium, referred to as the discovery cohort (11). PGS were calculated using a high-dimensional Bayesian regression framework, which includes a continuous shrinkage prior on the effect sizes of the included Single Nucleotide Polymorphisms (SNPs) (23). This method has the advantage that allows researchers to add all potential SNPs into the PGS, without clumping or choosing a p-value threshold to specify inclusion. This method has been found to be superior in comparison to other polygenic scoring methods, as it is able to explain the greatest amount of variance (23). Final PGS score included 754,458 SNPs.

As we consider different levels of exposure, and to ease interpretation, we categorized the distribution of PGS-BMI scores into quintals: Lowest, low, average, high, and highest risk. The mean and standard deviation of the PGS-BMI in each group are listed in Supplementary Table 1.

#### Mediators

When the children were about 10.7 years old, parents were asked to report on their parental feeding behavior using a questionnaire with a total of 13 items. Parents rated how commonly they engaged in different parental feeding behaviors. Exploratory factor analyses suggested three factors, with an eigenvalue >1. After oblique rotation, two items did not contribute sufficiently to any of the three factors (factors loadings <0.4), and where henceforth dropped. This final solution included three subscales (latent factors): Emotional feeding (4 items, example:” I cheer her up with something to eat if she is sad or upset”), Restriction (4 items, example: “I deliberately keep some foods out of her reach”), and Pressure to eat (3 items, example: “I insist that she eats all the food on the plate”). These three factors of parental feeding behavior are in line with the most studied constructs in the literature (48). Factors scores on these three parental feeding behaviors were considered as joint mediators between genetic liability and the outcome, BMI at 12 years, as previous interventions have taken a holistic approach aiming to modify a range of feeding behaviors instead of focusing on one specific one (49). A full list of items, response options frequencies, and subscales can be found in Supplementary Table 2.

#### Outcome

Height and weight were measured during clinic visits when the children were about 12 years old (mean□=□12.5 years, SD□=□0.6). Weight was measured with a Tanita Body Fat Analyzer (Tanita TBF UK Ltd) to the nearest 50□g. Height was measured to the nearest millimeter with the use of a Harpenden Stadiometer (Holtain Ltd). BMI was calculated by dividing weight (in kg) by height (in m) squared.

#### Covariates

High maternal education at birth of child was defined by mothers having completed education up to A-Levels, the requirement for applying to university in the UK. Additional covariates were sex of the child, as well as self-reported BMI of the mother prior to pregnancy.

### Analyses

We have adapted the interventional disparity measure approach of Micali et al (22). This method aims to estimate how much of the disparity in outcome (Y, BMI at 12 years) due to the difference in an exposure (X, PGS) remains after mediating factors (M, parental feeding) are modified by a potential intervention. In the context of genetic liability, this framework allows researchers to assess the magnitude of disparity that would remain if downstream factors were changed (21, 35). The diagram in Figure 3 illustrates this conceptual model.

The measure of interest (i.e. our target of estimation, or estimand) is defined as the interventional disparity direct effect (IDM-DE) which captures the disparity in outcome, due to being exposed versus not exposed to X that would be observed if we could intervene and set the mediator M to be distributed as if X was set to take the no exposure value (22). In our case, X, the PGS-BMI, has 5 levels (1=lowest risk, 2=lower risk, 3=average risk, 4=high risk, 5=highest risk), which we index by j. Hence, the IDM-DE is specified separately for *j*=2,3,4,5, with j=1 treated as the reference value. Specifically, let 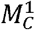 be a random draw from the distribution of M conditional on the confounder C when X is set to take the reference value 1, and Y(m) be the potential outcome when the mediator M is set to take the value m, in this case to take the randomly drawn value 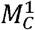. Note that M here is three-dimensional and therefore 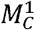 represents random draws from the joint distribution of the three parental behaviors.

The disparity measures of interest are then defined as, for j=2,3,4,5,

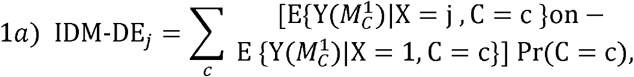

These four disparity measures capture the contrast between two levels of X while fixing the mediators to be distributed under a hypothetical scenario when X is set at the reference value 1. They represent the magnitude of the disparity in childhood BMI due to genetic liability (as captured by PGS) that would remain had all parental feeding behaviors been set at the lowest risk level (hypothetical intervention 1).

As this may be an unrealistic situation, we also defined these quantities for the hypothetical scenario where the reference distributions of parental behaviors, from which the random draws are taken, are those corresponding to the scenario where genetic liability is set at one risk category lower than the one they are observed to be in. For example, for a child in the highest risk (j=5) category, this hypothetical intervention would shift the distribution of parental feeding, as if they were in the high risk category (j=4). The same would be for the other categories, shifting from high risk (j=4) to average risk (j=3) and so on. For this setting, equation 1a is modified to allow for this shift in reference category, for j=2,3,4,5:

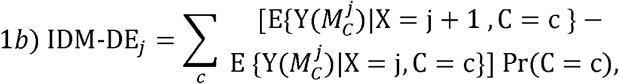

Like previous work (22), we consider interventions that change all mediators jointly, because it is unlikely that a hypothetical intervention addresses one parental feeding behavior only, as well as acknowledging that different aspects of parental feeding are likely to be correlated. Under the assumptions of no unmeasured confounding of the M-Y relationships, and of consistency for the mediators (i.e. that *E*{*Y*(*m*)|*X* = *j, C* = *c*} = *E*{*Y*|*X* = *j, C* = *c, M* = *m*}), as well as of no interference for the mediators, these quantities can be estimated from the data.

In addition, we also report estimates of the adjusted total association (Adj-TA) of PGS-BMI on BMI at 12 years, at each level of exposure (quintiles of genetic liability), in comparison to the referent (22). For j=1 treated as the reference group, i.e. hypothetical intervention 1, this is defined as, for j=2,3,4,5:

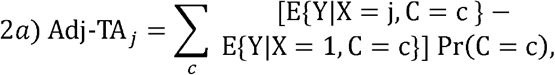

And for the hypothetical intervention 2, for j=2,3,4,5 this is amended:

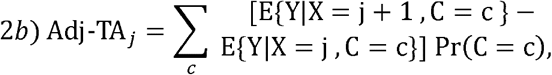

Analyses, consisting of a series of regressions for the mediators and outcome, were conducted in Stata version 16, with estimation carried out by plug-in parametric estimation and Monte Carlo simulation on a 1000-fold expanded dataset, with 1000 bootstrap samples. Regression models included interactions between confounders and mediators. The code is available on [https://github.com/MoritzHerle/Parental-feeding-and-childhood-genetic-risk-for-obesity] with details as in (22).

## Data Availability

Data are not available. For data access queries, please refer to: http://www.bristol.ac.uk/alspac/researchers/access/

## Acknowledgments

We are extremely grateful to all the families who took part in this study, the midwives for their help in recruiting them, and the whole ALSPAC team, which includes interviewers, computer and laboratory technicians, clerical workers, research scientists, volunteers, managers, receptionists, and nurses.

## Abbreviations

## Supplementary Information

**Supplementary Table 1.** Items measuring parental feeding practices when children were 10.7 years in ALSPAC

|  | Response options, N(%) |  |  |  |  |  |  |
| --- | --- | --- | --- | --- | --- | --- | --- |
| Items | Disagree | Slightly disagree | Neither agree or disagree | Slightly agree | Agree | Factor loadings | Subscale derived from factor analysis |
| I have to be sure that she does not eat too many sweets | 792 (11) | 296 (4) | 886 (12) | 2237 (30) | 3129 (43) | 0.53 | Restriction |
| I have to be sure that she does not eat too many of her favourite foods | 2103 (29) | 806 (11) | 2096 (29) | 1520 (21) | 788 (11) | 0.71 | Restriction |
| I deliberately keep some foods out of her reach | 4772 (65) | 414 (6) | 724 (10) | 748 (10) | 652 (9) | 0.67 | Restriction |
| It's OK to offer sweets as a reward for good behaviour | 1875 (26) | 945 (13) | 1738 (24) | 1741 (24) | 1030 (14) |  | Did not load on specific factor, and hence removed |
| If I did not guide or regulate her eating she would eat too much | 4369 (60) | 568 (8) | 686 (9) | 1025 (14) | 670 (9) | 0.7 | Restriction |
|  | **Never** |  | **Sometimes** |  | **Always** |  |  |
| I insist that she eats all the food on the plate | 2847 (41) |  | 3606 (51) |  | 572 (8) | 0.75 | Pressure to eat |
| If she does not finish all of the main course she is not allowed a pudding | 2487 (37) |  | 3401 (51) |  | 792 (12) | 0.73 | Pressure to eat |
| I tell her off for playing or fiddling with food at mealtimes | 2481 (39) |  | 3347 (53) |  | 538 (8) | 0.53 | Pressure to eat |
| I allow her to eat only at meal times, and not in between meals | 3200 (47) |  | 3408 (50) |  | 187 (3) |  | Did not load on specific factor, and hence removed |
| I cheer her up with something to eat if she is sad or upset | 3384 (50) |  | 3339 (49) |  | 79 (1) | 0.56 | Emotional feeding |
| I like to take her out for a special meal when something good happens to her | 1573 (23) |  | 5017 (73) |  | 317 (5) | 0.67 | Emotional feeding |
| I give her her favourite food when she is hurt or sick | 1414 (21) |  | 4748 (69) |  | 736 (11) | 0.71 | Emotional feeding |
| I like to prepare a special meal for her when something good happens to her | 1434 (21) |  | 5019 (73) |  | 462 (7) | 0.82 | Emotional feeding |

**Supplement Table 2.** Means and standard deviations of PGS-BMI across the 5 quintiles, n= 4,248

|  | **Mean** | **SD** |
| --- | --- | --- |
| **PGS-BMI** | -0.05 | 0.99 |
| **1<sup>st</sup> quintile**<br>**Lowest risk (N=849)** | -1.46 | 0.49 |
| **2<sup>nd</sup> quintile**<br>**Lower risk (N=850)** | -0.58 | 0.17 |
| **3<sup>rd</sup> quintile**<br>**Average risk (N=849)** | -0.04 | 0.14 |
| **4<sup>th</sup> quintile**<br>**Higher risk (N=850)** | 0.49 | 0.16 |
| **5<sup>th</sup> quintile**<br>**Highest risk (N=850)** | 1.33 | 0.43 |

Abbreviations: PGS-BMI = Polygenic Score BMI

**Supplementary Table 3.** Pairwise Pearson’s correlations between exposures, mediators, and outcome; n=4,248

|  | **PGS-BMI** | **Restriction** | **Emotional feeding** | **Pressure to eat** | **BMI at 12 years** |
| --- | --- | --- | --- | --- | --- |
| **PGS-BMI** | 1 |  |  |  |  |
| **Restriction** | 0.12 | 1 |  |  |  |
| **Emotional feeding** | 0.02 | 0.11 | 1 |  |  |
| **Pressure to eat** | 0 | -0.23 | -0.22 | 1 |  |
| **BMI at 12** | 0.36 | 0.3 | 0 | 0.1 | 1 |

Abbreviations: PGS-BMI = Polygenic Score BMI; BMI= Body Mass Index

